# Telehealth versus Self-Directed Lifestyle Intervention to Promote Healthy Blood Pressure: A Protocol for a Randomized Controlled Trial

**DOI:** 10.1101/2020.09.23.20200477

**Authors:** Mohamed Taher, Christina Yule, Heather Bonaparte, Sara Kwiecien, Charlotte Collins, Allison Naylor, Stephen P Juraschek, Lisa Bailey-Davis, Alex R Chang

## Abstract

**Introduction:** Weight loss, consumption of a Dietary Approaches to Stop Hypertension (DASH) dietary pattern, reduced sodium intake, and increased physical activity have been shown to lower blood pressure. Use of web-based tools and telehealth to deliver lifestyle counseling could be potentially scalable solutions to improve blood pressure through behavioral modification though limited data exists to support these approaches in clinical practice.

**Methods and analysis:** This randomized controlled trial will compare the efficacy of a telehealth versus self-directed lifestyle intervention in lowering 24-hour systolic blood pressure in patients with body mass index ≥ 25 kg/m^2^ and 24-hour SBP 120-160 mmHg. All participants receive personalized recommendations to improve dietary quality based on a web-based food frequency questionnaire, access to an online comprehensive weight management program, and a smartphone dietary app. The telehealth arm additionally includes weekly calls with registered dietitian nutritionists who use motivational interviewing. The primary outcome is change from baseline to 3-months in 24-hour systolic blood pressure. Secondary outcomes include changes from baseline in Healthy Eating Index-2015 score, weight, waist circumference, and physical activity.

**Ethics and dissemination:** The study has been approved by the Geisinger Institutional Review Board. Results will be disseminated through peer-reviewed publications and conference presentations.

**Trial registration number:** ClinicalTrials.gov Identifier NCT03700710

**WHO Trial Registration Data Set:**

1. Primary registry and trial identifying number: ClinicalTrials.gov Identifier NCT03700710
2. Date of registration in Primary Registry: 10/9/2018
3. Secondary identifying numbers: N/A
4. Source of Monetary or Material Support: Geisinger Health Plan
5. Primary Sponsor: Geisinger Clinic
6. Secondary Sponsors: None
7. Contact for Public Queries: Christina Yule; 570-214-5233;
8. Contact for Scientific Queries: Alex R. Chang, MD, MS; Kidney Health Research Institute; Geisinger; 100 N Academy Ave, Danville, PA; 570-271-8026;
9. Public title: Remote Dietary Counseling to Promote Healthy Diet and Blood Pressure
10. Scientific title: Remote Dietary Counseling to Promote Healthy Diet and Blood Pressure
11. Countries of recruitment: USA
12. Health conditions studied: high blood pressure
13. Interventions: <u>Active Comparator: Self-Guided Approach</u> In the self-guided arm, participants will receive access to web-based tools to help achieve healthy lifestyle changes to lower their blood pressure. The web-based tools include: 1) a web-based food frequency questionnaire (Viocare FFQ), which will provide a snapshot of participants’ dietary habits in the past 6 months as well as personalized recommendations for areas to improve; 2) access to BMIQ (www.bmiq.com), an evidence-based program developed by Dr. Louis Aronne at Columbia University, which includes program materials for weight loss and leading a healthy lifestyle; 3) LoseIt (www.loseit.com), a meal-logging app that integrates seamlessly with the BMIQ website. <u>Experimental: Dietitian-led Approach</u> In the dietitian-led arm, dietitian will use motivational interviewing in 15-30 minute telephone calls with participants. The BMIQ website will be used to share participant dietary data (LoseIt) and weight data with dietitians. Participants will receive access to web-based tools to help achieve healthy lifestyle changes to lower their blood pressure. The web-based tools include: 1) a web-based food frequency questionnaire (Viocare FFQ), which will provide a snapshot of participants’ dietary habits in the past 6 months as well as personalized recommendations for areas to improve; 2) access to BMIQ; 3) LoseIt (www.loseit.com), a meal-logging app that integrates seamlessly with the BMIQ website.
14. Key Inclusion and Exclusion Criteria Inclusion Criteria: 24-hour ambulatory SBP 120-160 mmHg, BMI ≥ 25 kg/m2, access to a telephone, access to a computer or smartphone with internet access, complete dietary data entry using LoseIt for at least 5 out of 7 days during run-in period, enter weight into the BMIQ portal during run-in period Exclusion Criteria: inability to understand English, myocardial infarction, stroke, or atherosclerotic cardiovascular disease procedure within last 6 months, current treatment for malignancy, planned or previous bariatric surgery, pregnant, breast-feeding, or planned pregnancy prior to the end of participation, self-reported average consumption of > 21 alcoholic beverages per week or binge drinking, psychiatric hospitalization in past year, current symptoms of angina, planning to leave the area prior to end of the study, current participation in another clinical trial, principal investigator discretion (i.e. concerns about safety, compliance)
15. Study type: randomized, parallel-arm, interventional study; study staff assessing outcomes and statistical analysis are masked. Purpose is to compare the efficacy of a self-guided vs. a telehealth dietitian-led approach to lower blood pressure through lifestyle modification.
16. Date of first enrollment – 1/16/19
17. Sample size – 200
18. Recruitment status – recruiting
19. Primary outcome - Change in 24-hour Systolic Blood Pressure [Time Frame: Baseline to 12-week follow-up] Measured by 24-hour ambulatory blood pressure monitoring (SpaceLabs Ontrak)
20. Key secondary outcomes Change in 24-hour diastolic blood pressure [Time Frame: Baseline to 12-week follow-up] Measured by 24-hour ambulatory blood pressure monitoring (SpaceLabs Ontrak) Change in Daytime Systolic Blood Pressure [Time Frame: Baseline to 12-week follow-up] Measured by 24-hour ambulatory blood pressure monitoring (SpaceLabs Ontrak) Change in Nighttime Systolic Blood Pressure [Time Frame: Baseline to 12-week follow-up] Measured by 24-hour ambulatory blood pressure monitoring (SpaceLabs Ontrak) Change in Daytime Diastolic Blood Pressure [Time Frame: Baseline to 12-week follow-up] Measured by 24-hour ambulatory blood pressure monitoring (SpaceLabs Ontrak) Change in Nighttime Diastolic Blood Pressure [Time Frame: Baseline to 12-week follow-up] Measured by 24-hour ambulatory blood pressure monitoring (SpaceLabs Ontrak) Change in Total Healthy Eating Index - 2015 score [Time Frame: Baseline to 12-week follow-up] Assessed by Viocare Food Frequency Questionnaire (score 0-100, 100=best possible score) Change in Weight [Time Frame: Baseline to 12-week follow-up] Weight measured at baseline and 12-week visits using a calibrated scale without shoes Change in Waist Circumference [Time Frame: Baseline to 12-week follow-up] Measured using Gulick II tape measure Change in Physical Activity (metabolic equivalent of task [MET]-minute per week [Time Frame: Baseline to 12-week follow-up] Measured by International Physical Activity Questionnaire (IPAQ) Short Form Change in Clinic systolic blood pressure (SBP) [Time Frame: Baseline to 12-week follow-up]Measured by average of 3 readings using Omron HEM 907XL Change in Clinic diastolic blood pressure (DBP) [Time Frame: Baseline to 12-week follow-up] Measured by average of 3 readings using Omron HEM 907XL
21. Ethics review – status approved, date of approval 10/2/2018
22. Completion date – ongoing
23. Summary results – N/A
24. IPD sharing statement – Deidentified data may be shared upon reasonable request.

**Article summary:** *Strengths and limitations of this study:*

- This randomized controlled trial will compare the efficacy of a telehealth vs. self-directed lifestyle intervention in lowering blood pressure through lifestyle modification in patients with elevated blood pressure and overweight/obesity
- The proposed interventions under investigation are low-cost and potentially scalable
- Primary endpoint data will be collected using 24-hour ambulatory blood pressure monitoring at baseline and at 3 months
- Additional secondary endpoint data will be collected including Healthy Eating Index-2015 score assessed by food frequency questionnaire, weight, waist circumference, and physical activity, assessed by questionnaire
- While participants and some of the study staff are unable to be blinded, researchers assessing study outcomes and conducting analyses will be blinded to the arms.

## Introduction

The ACC/AHA 2017 Hypertension Guidelines redefined hypertension as having 2 blood pressure readings measured on at least 2 separate occasions ≥ 130/80 mmHg(1). With this updated definition, nearly half of the U.S. adult population has hypertension(2). Obesity, Western-style dietary patterns rich in sodium and processed foods, and low physical activity have been implicated in the pathogenesis of hypertension and cardiovascular diseases (1, 3). Lifestyle modification is recommended for all patients with hypertension since weight reduction, consumption of a low-sodium Dietary Approaches to Stop Hypertension (DASH)-type diet, and increased physical activity can lower blood pressure substantially(4-6).

Despite recognition that addressing lifestyle behaviors is of high importance to address cardiometabolic risk, delivery of lifestyle modification in clinical care remains challenging due to time constraints, lack of referral resources, and other competing priorities(7, 8). Some possibilities to address this gap include the use of app-based interventions and the use of telehealth to deliver lifestyle counseling remotely. A recent systematic review and meta-analysis found that app-based interventions have the potential for aiding patients make healthy nutrition behavior changes and improving nutrition-related health outcomes (9). Prior studies have found that telehealth lifestyle coaching interventions are similarly effective as traditional, in-person delivered interventions for helping patients achieve weight loss(10, 11).

It remains unclear whether telehealth lifestyle counseling further lowers blood pressure through healthy behavior modification on top of app-based interventions. In this trial, we aim to compare the efficacy of a self-guided vs. a telehealth dietitian-led approach using web-based tools in patients with elevated blood pressure.

## Methods and analysis

### Study Design/setting

The Healthy BP study is a randomized controlled trial (RCT) comparing a self-guided vs. a dietitian-led telehealth approach to help patients lower blood pressure through lifestyle modification. The RCT is being conducted at 2 sites (Danville, PA, Wilkes-Barre, PA) within the Geisinger Health System. Recruitment began January 16, 2019.

### Study Population

A total of 200 participants will be included. Eligibility criteria include ≥ 18 years of age, body mass index (BMI) > 25 kg/m^2^, access to a telephone and either a computer or smartphone with internet access, 24-hour ambulatory systolic blood pressure between 120-160 mmHg, and successful completion of the run-in period [enter dietary data entry into LoseIt app (www.loseit.com) for at least 5 out of 7 days; enter weight into a comprehensive weight management website (www.bmiq.com)]. Exclusion criteria include: inability to understand English, myocardial infarction, stroke, or atherosclerotic cardiovascular disease within prior 6 months, planned or previous bariatric surgery, pregnancy, breast-feeding, or planned pregnancy prior to the end of participation, self-reported average consumption of > 21 alcoholic beverages per week or binge drinking, psychiatric hospitalization in past year, current angina, or plans to leave the area prior to the end of the study, current participation in another clinical trial, and principal investigator discretion (i.e. concerns about safety, compliance).

### Interventions

All participants in both arms will receive instructions to access the BMIQ website and set up an account on LoseIt, a meal-logging app that integrates with the BMIQ website. BMIQ is an evidence-based, Health Insurance Portability and Accountability Act (HIPAA)-compliant program developed by Dr. Louis Aronne(12). Educational materials on the BMIQ website include materials to promote weight loss, eating a DASH-type diet (increasing intake of fruits/vegetables, whole grains, protein sources from plants, lean meats, and seafood, and decrease sodium, sugar intake, and saturated fat), reducing sodium intake, increasing physical activity, setting goals, overcoming barriers, and relapse prevention. A list of the session topics, with a few customizations by the study team, are shown in **Table 1**. Participants also will receive a personalized nutrition report based on the Viocare Food Frequency Questionnaire (FFQ) that suggest ways to improve their dietary habits to best align with national dietary guidelines(13). If participants encounter problems with using LoseIt or BMIQ, research staff will provide assistance. If no data is logged for more than a week, research staff will reach out to patients via text messages or phone calls to help troubleshoot any problems. To encourage adherence with dietary data entry, participants will receive virtual lottery tickets (lottery drawings for ten $300 gift certificates) each week in which they enter at least 5 days/week of dietary data and 2 days/week of weights. All participants will be offered the opportunity to participate in grocery store tours at a local grocery store (Weis Markets).

**Table 1.**
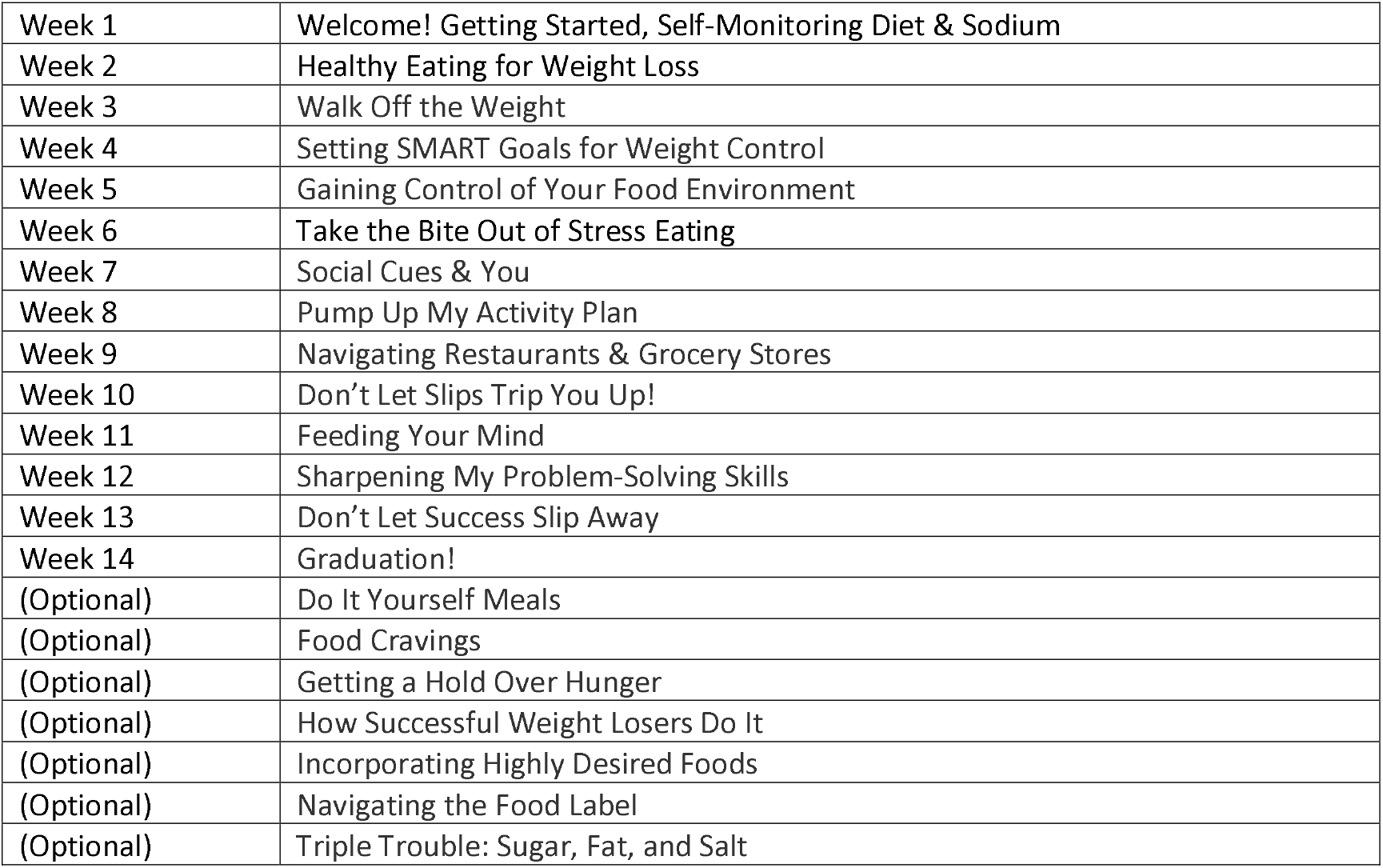
BMIQ Session Topics.

|  |  |
| --- | --- |
| Week 1 | Welcome! Getting Started, Self-Monitoring Diet & Sodium |
| Week 2 | Healthy Eating for Weight Loss |
| Week 3 | Walk Off the Weight |
| Week 4 | Setting SMART Goals for Weight Control |
| Week 5 | Gaining Control of Your Food Environment |
| Week 6 | Take the Bite Out of Stress Eating |
| Week 7 | Social Cues & You |
| Week 8 | Pump Up My Activity Plan |
| Week 9 | Navigating Restaurants & Grocery Stores |
| Week 10 | Don't Let Slips Trip You Up! |
| Week 11 | Feeding Your Mind |
| Week 12 | Sharpening My Problem-Solving Skills |
| Week 13 | Don't Let Success Slip Away |
| Week 14 | Graduation! |
| (Optional) | Do It Yourself Meals |
| (Optional) | Food Cravings |
| (Optional) | Getting a Hold Over Hunger |
| (Optional) | How Successful Weight Losers Do It |
| (Optional) | Incorporating Highly Desired Foods |
| (Optional) | Navigating the Food Label |
| (Optional) | Triple Trouble: Sugar, Fat, and Salt |

Overarching participant goals include: 1) weight loss of 3% at 3 months; 2) consume a healthier dietary pattern (high in fruits, vegetables, whole grains, low-fat dairy, vegetable/fish/poultry sources of protein, healthier sources of fat, and avoid sugar and salt); 3) reduce sodium intake to <2300 mg/d; 4) at least 180 min/wk of moderate/vigorous intensity physical activity.

<u>The self-guided intervention group</u> will receive access to web materials but no additional support from dietitians.

<u>The telehealth dietitian-led intervention group</u> additionally will receive weekly telephone calls (initial call 30-60 minutes, follow-up calls 15-20 minutes) from a study dietitian. This approach is informed by Carver and Scheier’s control systems theory(14) and the situated learning theoretical framework(15). Control systems theory describes human behavior as a “continual process of moving toward, and away from, various kinds of mental goal representations, and this movement occurs by a process of feedback control.” By using the app and website and conversing with dietitians remotely, situated learning will occur through activities and experiences in the context of participants’ own environments. Dietitians in this study explore participants’ goals and values and help patients set an attainable goal around a health behavior for the week and assess their progress towards those goals using motivational interviewing techniques and principles from. Motivational interviewing skills will be continually developed through bi-annual training sessions by a motivational interviewing expert. Dietary data entered in the LoseIt app is used to estimate average daily intake of sodium, fruits, and non-starchy vegetables, which can be used for guidance for goal-setting around sodium intake and fruit and vegetable intake.

### Recruitment and Baseline 24-hour ABPM

Participants will be recruited using multiple methods. The primary source of recruitment will be done by identifying patients with elevated outpatient blood pressure using Geisinger electronic health record (EHR) data. Additional sources of recruitment will include patients with Geisinger Health Plan insurance who have elevated blood pressure during health screenings and referrals from Geisinger providers and self-referrals. Potential participants will then be invited to complete a baseline 24-hour ambulatory blood pressure monitoring (ABPM) to confirm their elevated blood pressure outside the office as recommended by guidelines(16). The 24-hr ABPM will be provided to participants free of charge, using SpaceLabs Ontrak devices. Participants will be asked to retake the 24-hour ABPM if fewer than 14 daytime measurements or 7 nighttime measurements are obtained. Awake and sleep times will be self-reported; if participants fail to report their awake and sleep times on the activity diary, the SpaceLabs Ontrak activity monitor will be examined visually to determine awake and sleep times(17). The study flow is shown in **Figure 1**.

**Figure 1.**
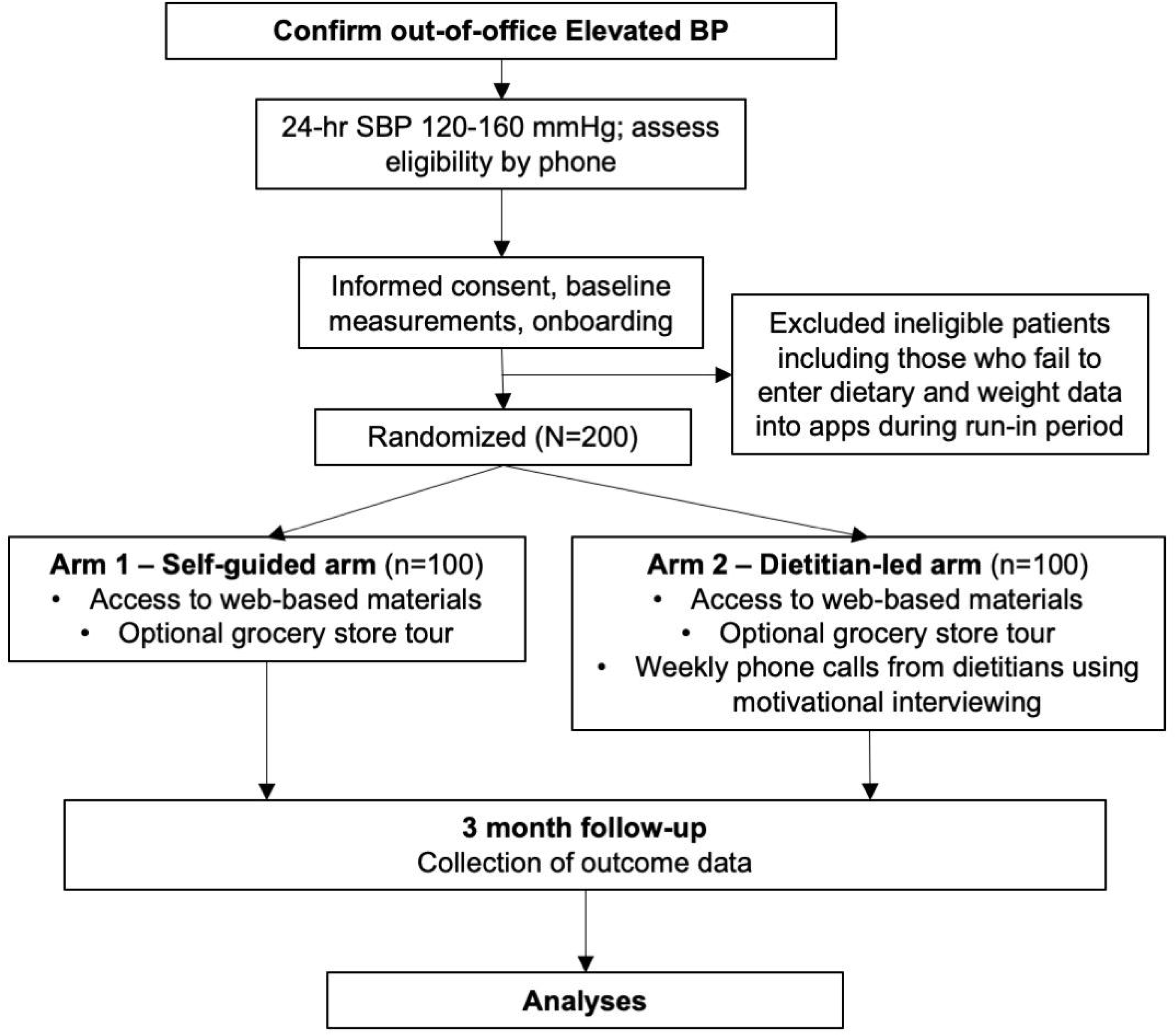
Study Flow.

### Baseline Study Visit, Run-in period, Randomization and Blinding

Participants who meet inclusion criteria are then invited to an initial study visit where informed consent will be obtained, along with weight, waist circumference, and completion of Viocare FFQ(13), the International Physical Activity Questionnaire (IPAQ) Short Form(18), and other study questionnaires.

Participants will register for a BMIQ web account, download the LoseIt app, and shown how to sync LoseIt to BMIQ, and access various features of LoseIt and BMIQ. Participants will then be instructed to enter dietary data daily and weight data during the run-in period and throughout the trial. If participants enter complete dietary data (3 eating occasions per day) for 5/7 days and enter their weight into the BMIQ website, they will be eligible to continue to randomization.

Participants who are struggling with completing dietary data entry during the run-in period will be contacted by telephone to inquire about any issues, and the run-in period will be extended until at least 5 out of 7 consecutive days of dietary data entry is complete. Participants who complete the run-in period will then by contacted by telephone for randomization assignment. Randomization assignments will be generated by a computer program, and an unblinded staff member will open a sealed opaque envelope with the randomized intervention sequence, and notify participants of their intervention group assignment.

Participants, dietitians, and the member of the study team assigning randomization cannot be blinded. All other members of the study team are blinded, including study team members assessing outcomes and conducting statistical analyses.

### Outcome Assessment

Outcomes will be assessed as change from baseline to the 3-month follow-up visit. The primary outcome is change in 24-hour systolic blood pressure, assessed using ambulatory blood pressure monitoring (ABPM) with SpaceLabs Ontrak ABPM devices(17). Secondary outcomes include changes in total Healthy Eating Index (HEI)-2015 score, weight, waist circumference, 24-hour diastolic blood pressure, clinic systolic blood pressure, clinic diastolic blood pressure, and physical activity (total metabolic equivalent of task [MET]-minutes per week). The HEI-2015 score will be assessed using the Viocare FFQ(19), and total MET-minutes per week assessed using the IPAQ-short form. Weight will be measured without shoes or outerwear using a calibrated scale in clinic or by home scales—in the case of remote research visits necessitated by the Novel Coronavirus Disease 2019 (COVID-19) pandemic (**Table 2**). Clinic blood pressure is measured as the average of 3 measurements taken 1-minute apart after a 5-minute rest using the Omron 907-XL device. Other outcomes include change in individual components of the HEI-2015 score and patient satisfaction (5-point Likert score).

**Table 2.** Adaptations to trial during COVID-19.

|  |  |
| --- | --- |
| Recruitment | Research personnel unable to conduct in-person visits from March 16, 2020-current. Recruitment reliant on referrals. |
| Baseline 24-hr ABPM | Increased collaboration with clinical staff to perform 24-hr ABPM testing for patients with elevated outpatient blood pressure readings |
| Baseline study visit | Study visits planned for late March 2020 were postponed until remote adaptations were made and approved by the IRB. Remote baseline study visits began in May 2020 using telephone calls and study materials mailed or dropped off at participants' houses. Weight and waist circumference measured using home scales and tape measures (rather than in clinic); automated office blood pressure unable to be performed. |
| Grocery store tour | Weis Markets introduced a virtual grocery store tour option for participants |
| Follow-up visit and 24-hr ABPM | Follow-up study visits and 24-hr ABPMs in late March 2020 were postponed until remote adaptations were made and approved by the IRB. Remote follow-up visits and 24-hr ABPMs were conducted using telephone calls and study materials mailed or dropped off at participants' houses. Weight and waist circumference measured using home scales and tape measures (rather than in clinic); clinic automated office blood pressure unable to be performed. |

### Adaptations to trial during COVID-19 pandemic

In late March 2020 and onwards, adaptations were made due to the COVID-19 pandemic, which necessitated minimizing in-person contacts to preserve clinic space to allow for social distancing and conserving personal protective equipment, (**Table 2**). Fortunately, we were able to continue the trial with adaptations although time windows were necessarily extended by approximately 1 month during March and April 2020 as there was a large degree of uncertainty on how to proceed and stay-at-home orders in Pennsylvania were in effect.

### Sample size calculation

The original sample size plan was for 160 patients, based on a conservative estimated drop-out rate of 25% and a standard deviation (SD) of 8.8 mmHg change in 24-hour SBP to detect a difference of 4.6 mmHg between study arms. In order to determine whether there was a need to request additional funding from the funder (Geisinger Health Plan), a preplanned interim analysis was performed on 9/13/19, showing that the SD for change in 24-hour SBP was larger than expected (SD 9.9) in the first 28 participants. Thus, a revised power calculation was performed. With an anticipated drop-out rate of 25%, the updated sample size of 200 patients (150 completers) we expect to have >80% power to detect a difference of 4.6 mmHg between study arms in change of 24-hour SBP. For other outcomes, we expect to have >80% power to detect a difference of 6.9 in total HEI-2015 score with a SD of 15 units for change in total HEI-2015 score, and >80% power to detect a difference of 2.1 kg between groups, assuming a SD of 4.5 kg for change in weight.

### Statistical analysis

The primary contrast will be the difference from baseline between treatment group. The primary analyses will be intention-to-treat including all randomized participants who complete the trial with outcomes data, using unpaired t tests. We will also examine whether 24-hour systolic blood pressure improves at 12 weeks compared to baseline for each arm. We will use a 2-sided alpha of 0.05. Sensitivity analyses excluding non-compliant patients (< 25% days with complete dietary data) will also be conducted. In addition, in sensitivity analyses we will compare the end of period values (not baseline change) with and without adjustment for baseline. We will test for an interaction between treatment assignment and baseline 24-hour systolic blood pressure (≥ or < 130 mmHg) as an analysis of the DASH-Sodium trial found that low-sodium intake and the DASH diet had greater BP-lowering effects in those with higher baseline systolic BP(20). All outcome measures will be assessed for a normal distribution, and non-normally distributed outcomes will be log-transformed. Likert scales will be assessed on a continuous scale or dichotomized. Continuous outcomes will be compared via linear regression, while dichotomized outcomes will be compared via logistic regression.

### Ethics and dissemination

Initial ethical review and approval for this study was obtained from the Geisinger Institutional Review Board (IRB) on 10/2/2018 (current protocol version 1.22; approved 8/6/2020). Protocol amendments will be communicated to members of the study team and updated on the clinicaltrials.gov website. Data will be collected using REDCap software on a secure, password-protected server. Study investigators are responsible for ensuring adherence to the protocol and guidelines for Good Clinical Practice. Symptomatic hypotension is possible in patients using blood pressure-lowering medications and hypoglycemia is possible for patients on diabetes medications undergoing dietary changes. Participants will be educated about the symptoms of hypotension and hypoglycemia. Study staff will inform investigators of all patient adverse events occurring throughout the trial.

Dissemination of research will occur through several pathways. Research results will be submitted for publication to peer-reviewed journals and presented at national conferences. Summaries of Healthy BP findings will be provided locally to Geisinger leadership and healthcare providers and nationally to other stakeholders and through press releases. After completing the study, participants and their primary care providers receive a summary of study results.

### Patient and public involvement

There was no specific patient or public involvement in the planning of this study. We will disseminate summary findings from this trial to study participants and to Geisinger’s patient population.

## Data Availability

Deidentified data may be available upon reasonable request.

## Authors’ contributions

AC and LBD developed the idea for the study and obtained funding, All authors contributed to the protocol development. MT wrote the initial draft supervised by AC, and all authors have approved the final version.

## Acknowledgments

A version of this work was presented at the American Heart Association EPI/Lifestyle 2020 meeting March 3, 2020 in Phoenix, Arizona. We thank Dr. Louis Aronne and Guadalupe Minero for their guidance in setting up and customizing the BMIQ online platform.

